## Supplementa Table 1 for "Prevalence, severity, and risk factors of disability among adults living with HIV accessing routine outpatient HIV care in London, United Kingdom (UK): A cross-sectional self-report study"

**Supplemental File Table 1: WHODAS response per item, with Mean and Median scores per item (n=200)**

| **WHODAS items of functional limitation (disability domain)** | **No difficulty,**  **n (%)** | **Mild difficulty,**  **n (%)** | **Moderate difficulty,**  **n (%)** | **Severe difficulty,**  **n (%)** | **Extreme difficulty**  **or cannot do, n (%)** | **Item score,**  **Mean (SD);**  **Median (LQ, UQ)** |
| --- | --- | --- | --- | --- | --- | --- |
| Standing for long periods  (mobility domain) | 127 (63.5) | 24 (12.0) | 27 (13.5) | 12 (6.0) | 10 (5.0) | 0.8 (1.2);  0.0 (0.0, 1.0) |
| Household responsibilities  (life activities domain) | 121 (60.5) | 22 (11.0) | 34 (17.0) | 15 (4.0) | 8 (4.0) | 0.8 (1.2);  0.0 (0.0, 2.0) |
| Learning a new task  (cognition domain) | 145 (72.5) | 21 (10.5) | 22 (11.0) | 3 (1.5) | 9 (4.5) | 0.6 (1.1);  0.0 (0.0, 2.0) |
| Joining in community activities  (participation domain) | 123 (61.5) | 22 (11.0) | 24 (12.0) | 14 (7.0) | 17 (8.5) | 0.9 (1.3);  0.0 (0.0, 2.0) |
| Emotionally affected by health problems (participation domain) | 69 (34.5) | 43 (21.5) | 44 (22.0) | 31 (15.5) | 13 (6.5) | 1.4 (1.3);  0.0 (0.0, 2.0) |
| Concentrating for 10 minutes  (cognition domain) | 108 (54.0) | 45 (22.5) | 27 (22.5) | 14 (7.0) | 6 (3.0) | 0.8 (1.1);  0.0 (0.0, 2.0) |
| Walking a long distance  (mobility domain) | 128 (64.0) | 25 (12.5) | 19 (9.5) | 8 (4.0) | 20 (10.0) | 0.8 (1.3);  0.0 (0.0, 1.0) |
| Washing whole body  (self-care domain) | 156 (78.0) | 20 (10.0) | 11 (5.5) | 6 (3.0) | 7 (3.5) | 0.4 (1.0);  0.0 (0.0, 0.0) |
| Getting dressed  (self-care domain) | 154 (77.0) | 17 (8.5) | 20 (10.0) | 7 (3.5) | 2 (1.0) | 0.4 (0.9);  0.0 (0.0, 0.0) |
| Dealing with people you do not know  (getting along domain) | 121 (60.5) | 26 (13.0) | 36 (18.0) | 12 (6.0) | 5 (2.5) | 0.8 (1.1);  0.0 (0.0, 2.0) |
| Maintaining a friendship  (getting along domain) | 129 (64.5) | 29 (14.5) | 23 (11.5) | 12 (6.0) | 7 (3.5) | 0.7 (1.1);  0.0 (0.0, 1.0) |
| Day-to-day work  (life activities domain) | 113 (56.5) | 39 (19.5) | 18 (9.0) | 9 (4.5) | 21 (10.5) | 0.9 (1.3);  0.0 (0.0, 1.0) |

SD: Standard Deviation

LQ: Lower Quartile

UQ: Upper Quartile
